# Redefining Autism Subtypes: a machine learning approach leveraging topological data analysis, network measures and hemispheric lateralization

**DOI:** 10.1101/2025.04.14.25325816

**Authors:** Caroline L. Alves, Loriz Francisco Sallum, Patrícia Maria de Carvalho Aguiar, Joel Augusto Moura Porto, Francisco Aparecido Rodrigues, Thaise G. L. de O. Toutain, Michael Moeckel

## Abstract

Autism subtypes, including general Autism Spectrum Disorder (ASD) and Asperger Syndrome (AS), exhibit distinct neural connectivity patterns. This study is the first to systematically integrate Topological Data Analysis (TDA) with complex network measures and machine learning (ML) to investigate brain lateralization and connectivity differences among these subtypes. Using fMRI-derived connectivity matrices, TDA metrics—such as persistence entropy and fractal dimension—revealed that AS networks are highly integrated and hierarchically complex, distinguishing them from both ASD and typically developing (TD) groups. Shapley Additive Explanations (SHAP) analysis identified the left primary motor cortex as a key feature across all subtypes, and highlighted its subtype-specific correlations with other brain regions. ML models trained on these features achieved high classification accuracy, with an AUC of 0.983. This fMRI-based analysis supports the classification of AS as a distinct group alongside ASD due to its unique neurobiological characteristics.

## 1 Introduction

Autism Spectrum Disorder (ASD) is a neurodevelopmental condition with complex genetic underpinnings and significant heterogeneity, as highlighted by extensive research in the field [1, 2]. ASD typically manifests within the first three years of life and is characterized by challenges in social communication as well as restricted and repetitive behaviors [3]. Historically, subtypes like Pervasive Developmental Disorder-Not Otherwise Specified (PDD-NOS) and Asperger Syndrome (AS) were recognized under the Diagnostic and Statistical Manual of Mental Disorders, Fourth Edition (DSM-IV) [4, 5]. However, the updated DSM-V framework unified these subtypes under a single ASD diagnosis, emphasizing a spectrum of symptom presentations rather than distinct categories [6, 7].

The DSM-V’s shift toward a unified ASD diagnosis has sparked debate. Proponents argue that it simplifies diagnostic criteria, reflecting the overlapping features among subtypes, such as shared deficits in visuospatial and executive functions [8]. Critics, however, highlight that significant differences exist in the neural underpinnings, cognitive profiles, and developmental trajectories of subtypes like AS and PDD-NOS [9]. For instance, PDD-NOS is associated with difficulties in social relationships, communication, and motor skills [10, 11], whereas AS presents distinct characteristics, including narrow interests, repetitive activities, and the absence of significant language delays, often classifying it as a ”high-functioning” form of ASD [5, 12]. These differences raise the question: Does the DSM-V’s unified approach sufficiently capture the nuanced neurobiological and behavioral distinctions, or does the DSM-IV’s separation of subtypes provide a more precise framework for understanding and treating these conditions?

While substantial work has explored binary classification between ASD and typically developing (TD) individuals [13–15], there is a notable gap in research focused on identifying distinct ASD subtypes using advanced methodologies. Few studies have investigated subtype classification using machine learning (ML). For instance, convolutional neural networks (CNNs) have been applied to resting-state fMRI data, achieving accuracy of 0.821 for DSM-IV-defined subtypes [16], while other ML models yielded similar performance using behavioral checklists [17, 18]. However, these efforts often lack detailed biological interpretability and do not leverage advanced tools like topological data analysis (TDA) or community detection metrics to elucidate the structural and functional distinctions among subtypes.

In this study, we aim to address these gaps. We evaluate whether evidence based on biological data supports the distinctions proposed by the DSM-IV and whether they still hold clinical and scientific relevance. Specifically, we investigate brain connectivity differences across ASD, AS, PDD-NOS, and TD groups using functional Magnetic Resonance Imaging (fMRI) data from the Autism Brain Imaging Data Exchange (ABIDE) [19], which leverages blood oxygenation level-dependent (BOLD) time series data. Our methodology introduces novel aspects, including the application of TDA to explore higher-order connectivity, using a systematic set of 26 complex network measures, and integrating ML techniques with Shapley Additive Explanations (SHAP) values for interpretability. TDA metrics, such as persistence entropy and fractal dimension, provide insights into the hierarchical and multi-scale topology of brain networks [20, 21]. Additionally, we analyze hemispheric differences to investigate lateralization patterns, motivated by evidence of atypical hemispheric specialization in ASD [22, 23].

By systematically comparing global and regional connectivity patterns across subtypes, we seek to determine whether the distinctions between AS, PDD-NOS, and ASD are biologically justified or whether the DSM-V’s unified approach better captures the complexity of these conditions. Our findings may have significant implications for refining diagnostic criteria, improving targeted interventions, and advancing our understanding of the neurobiological basis of ASD subtypes.

In summary, this study presents the first comprehensive framework integrating TDA, complex network measures, and ML to evaluate ASD subtypes. By uncovering nuanced distinctions in brain network topology, we aim to revisit the ongoing debate about whether separating subtypes, as in the DSM-IV, offers greater insight into the neural mechanisms of ASD than the unified approach of the DSM-V.

## 2 Results

### 2.1 Summary of the main results

This study provides novel insights into the neurobiological underpinnings of ASD subtypes, focusing on distinct connectivity patterns across TD (N=518), ASD (N=282), AS (N=80), and PDD-NOS (N=48). The data from the ABIDE dataset included BOLD time series from 122 brain regions, with preprocessing details described in sub-section 4.1. Our methodology employed ML classifiers together with advanced metrics such as complex network and TDA measures.

Our analysis highlighted the left primary motor cortex (Left-PrimMotor) as central to all subtypes, with additional group-specific connections identified, such as those involving the left orbitofrontal cortex (Left-OrbFrontal) in ASD and the left temporal pole (Left-Temporalpole) in AS. Cluster analysis revealed that PDD-NOS is closely related to ASD, while AS emerged as more distinct, suggesting nuanced differences within the spectrum (results detailed in Subsection 2.3).

Key novel findings include:

- **Benchmarking ML models for subtype classification:** ML results show clear separation of AS, while ASD exhibits less distinct patterns with other subtypes, indicating heterogeneity within ASD classifications (results detailed in Subsection 2.2);
- **Spatiotemporal correlation patterns:** Identification of the most relevant inter-regional brain correlations for each group, revealing unique functional connectivity signatures that differentiate AS, ASD, and TD (results detailed in Subsection 2.3);
- **Complex network measures highlight topological alterations in AS:** AS networks exhibit enhanced long-range connectivity and stronger local clustering, traits often associated with high-functioning autism (results detailed in Subsection 2.4).
- **Unique topological features in AS:** TDA metrics, including the fractal dimension and persistence entropy, revealed greater network complexity and hierarchical organization in AS. The elevated fractal dimension and topological feature variability underscore this subtype’s distinct neural connectivity patterns (results detailed in Subsection 2.5).
- **Hemispheric and regional insights:** AS networks demonstrated pronounced hemispheric asymmetries, with reduced right-hemisphere efficiency and larger left-hemisphere diameter compared to ASD and TD. These findings highlight the interplay between localized inefficiencies and global robustness in AS (results detailed in Subsection 2.6).

### 2.2 Benchmarking ML models for subtype classification

In order to conduct our study, a diverse set of machine learning classifiers was used, including the support vector machine (SVM) [24], logistic regression (LR) [25] employing the limited-memory Broyden Fletcher Goldfarb Shanno (L-BFGS) solver, random forest (RF) [26], multilayer perceptron (MLP) [27], and convolutional neural network (CNN) [28, 29]. For evaluation, we used accuracy as the primary metric to assess the overall correctness of the classification model [30–34], along with precision and recall to evaluate class-specific performance [35–38]. Precision quantifies the model’s ability to correctly classify instances within specific categories like TD, while recall (sensitivity) assesses its effectiveness in identifying positive examples across ASD, AS, and PDD-NOS. To visualize performance, we used receiver operating characteristic (ROC) curves, with the area under the ROC curve (AUC) serving as a key metric, ranging from 0.5 (random performance) to 1 (perfect classification) [30, 39–41]. We calculated the micro-average AUC, which treats all classes equally, and the macro-average AUC, which aggregates the contributions of each class individually, to assess both individual and overall classification performance [42].

According to Figure 1, the best classifiers were CNN and SVM. For the test set, CNN achieved a mean AUC of 0.983, precision of 0.962, recall of 0.962, and accuracy of 0.981. On the other hand, SVM achieved an AUC of 0.962, precision of 0.989, recall of 0.928, and accuracy of 0.986.

**Fig. 1.**
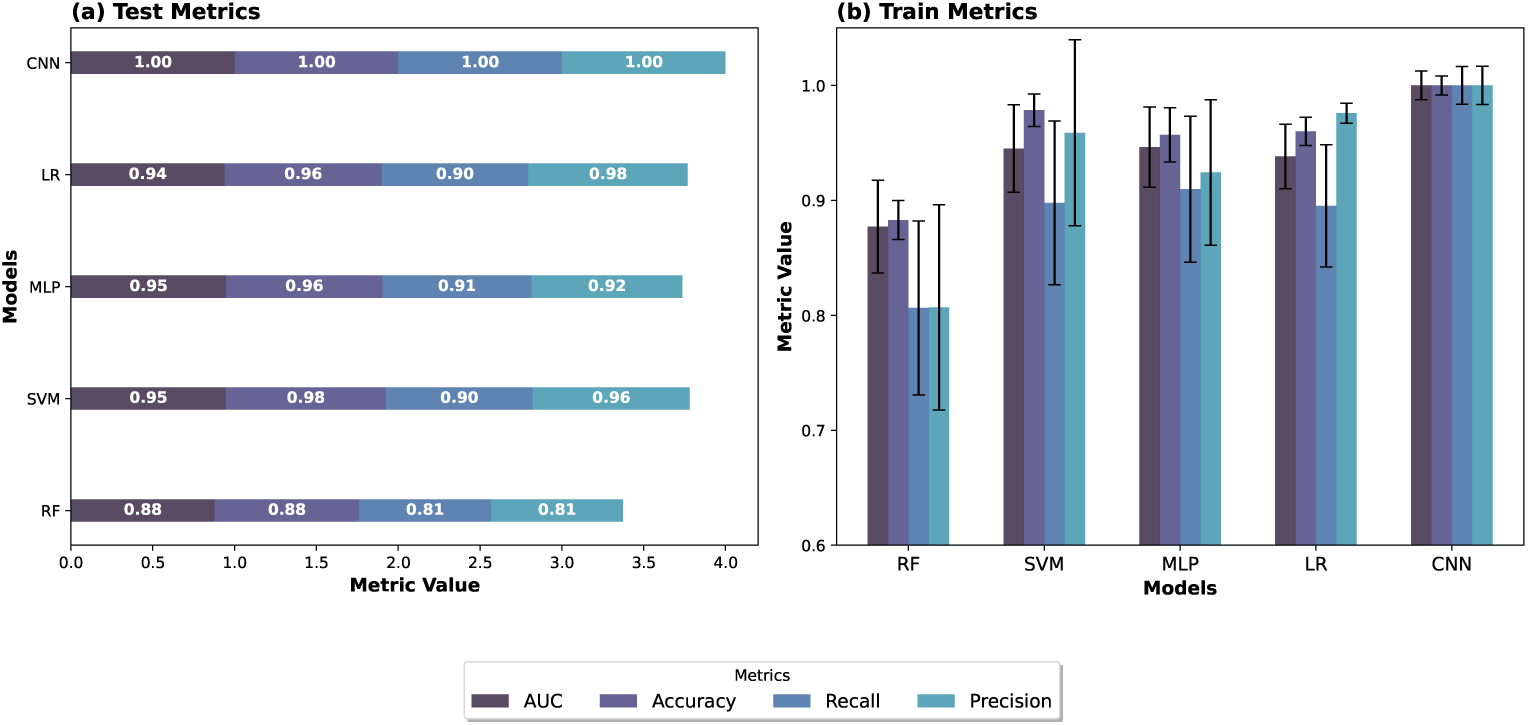
Results of all ML classifiers: (a) the results for the test set and (b) the results for the training set, with error bars corresponding to the 10-fold stratified cross-validation. The best performance was achieved by CNN and SVM.

Since SVM has a lower computational cost, it was chosen for the subsequent steps. Figure 2 displays the confusion matrix (Figure 2-(a)), the learning curve (Figure 2-(b)), the ROC curve (2-(c)), and the precision-recall curve ((2-(c))), respectively. From Figures 2-(a) and (b), the confusion matrix and ROC curve, respectively, show that the group most difficult to distinguish was PDD-NOS (in the confusion matrix, PDD-NOS group has some mistaken classification with ASD group). A possible explanation for this is that PDD-NOS is indeed a subclass of ASD, leading to overlapping features between the groups. Additionally, this group has the least amount of data, which could potentially contribute to higher misclassification rates. In contrast, AS, ASD, and TD groups were clearly separated, suggesting that AS could be distinguished from

**Fig. 2.**
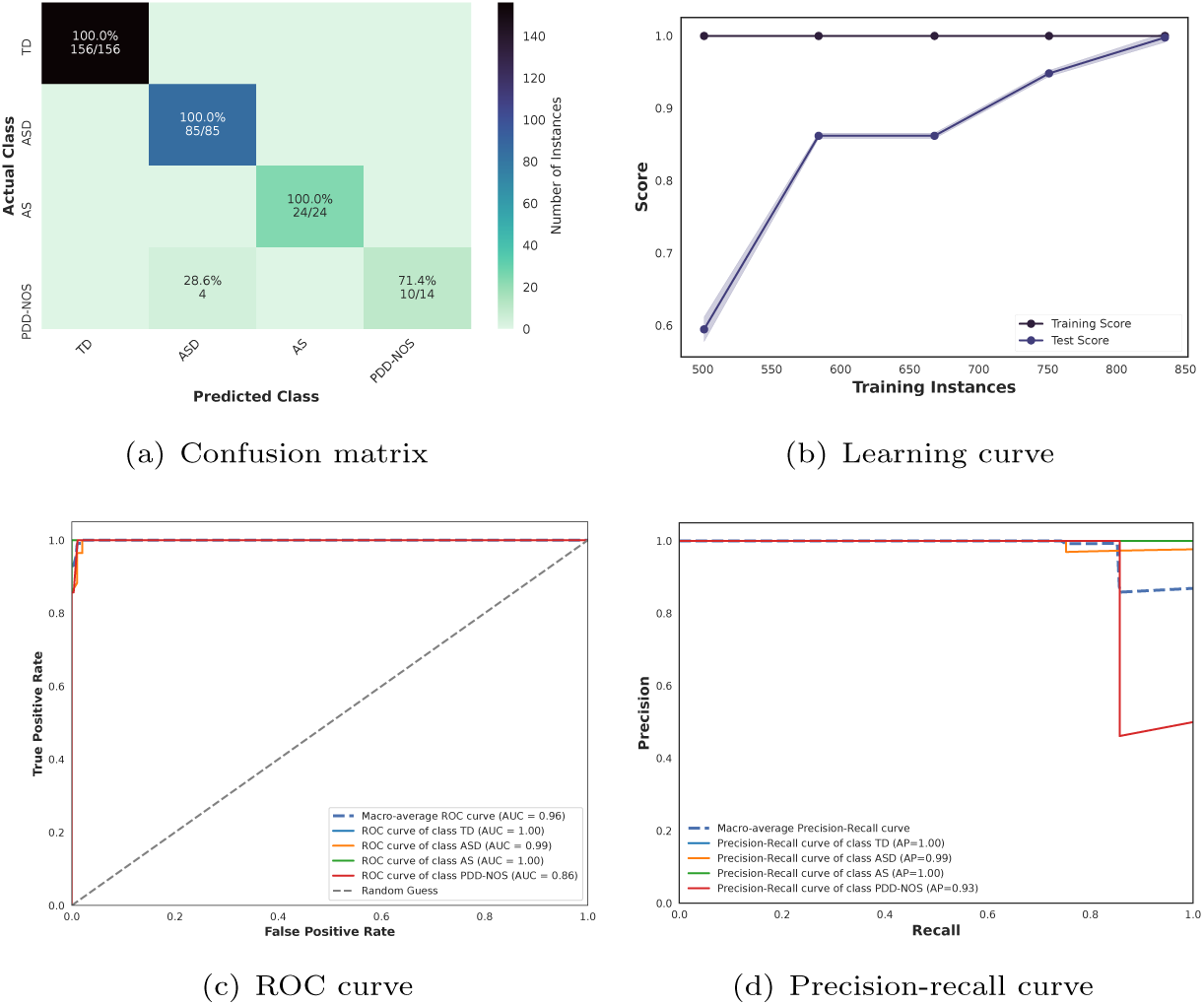
ML results on test sample from connectivity matrices. (a) Confusion Matrix depicting the performance of various ML algorithms on a test sample. The diagonal elements represent true positives (TP) values, indicating each algorithm’s accuracy in correctly identifying positive instances. (b) Learning curve showing the training accuracy (purple) and test accuracy (blue) across different training sizes. (c) ROC curve for each class. The dashed grey line represents the random choice classifier, while the dashed blue line denotes the macro-average ROC curve. The solid blue, orange, green, and red lines correspond to the ROC curves for the TD, ASD, AS, and PDD-NOS classes, respectively. (d) Precision-Recall (PR) curve for each class. The blue dashed line represents the macro-average PR curve, while the solid blue, orange, green, and red lines correspond to the PR curves for the TD, ASD, AS, and PDD-NOS classes, respectively.

ASD group, further supporting its classification as a distinct subtype within the ASD spectrum.

The visual representation of the learning curve illustrates the impact of varying the number of training instances on the model’s predictive accuracy [43]. Figure 2-(c) shows that the model requires the full dataset to achieve convergence. To address class imbalance due to differing numbers of patients in each diagnostic group, we computed Precision-Recall (PR) curves. These curves provide a comprehensive assessment of the model’s performance by depicting the trade-off between precision (the proportion of true positive predictions among all positive predictions) and recall (the proportion of true positives correctly identified) [44]. As shown in Figure 2-(d), the macro-average PR curve summarizes the overall performance across all classes, while individual curves capture class-specific behavior. Despite the class imbalance, particularly the smaller sample size of PDD-NOS group, the PR curves indicate that the model maintains robust classification performance across all groups. The results reveal high Average Precision (AP) for the TD (1.00), ASD (0.99), and AS (1.00) classes, underscoring the model’s ability to accurately distinguish these groups. Although PDD-NOS class exhibited a slightly lower AP (0.93), likely due to its limited sample size and feature overlap with the other groups, the overall macro-average performance highlights the model’s effectiveness in handling the imbalanced dataset and maintaining high classification accuracy.

### 2.3 Spatiotemporal correlation patterns

The methodology used for SHAP value analysis produced the results shown in Figure 12 in Appendix A, which highlights the primary region involved in all connections across different autism subtypes. Notably, the Left-PrimMotor region consistently emerged as the central region for all subtypes. Specifically, for the ASD group, the predominant connections included Left-PrimMotor and Left-OrbFrontal, followed by Outside Brodmann Area 5 (Outside BAS5) and Left-PrimMotor. In the AS group, the primary connections were Left-PrimMotor and Left-Temporalpole, followed by Outside Brodmann Area 1 (Outside BAS1) and Left-PrimMotor. Similarly, for the PDD-NOS group, the key connections comprised Left-PrimMotor and Left Thalamus (Left-Thalamus), as well as Left Secondary Visual Cortex (Left-SecVisual) and Left-PrimMotor. To identify in more detail the Outside BAS regions that played a key role in these correlations, we used the Yale BioImage Suite Package web application to visualize these areas, as shown in Figure 3. Based on [45, 46], we identified Outside BAS5 (Figure 3-(b)) as the posterior cerebellum, and Outside BAS1 (Figure 3-(a)) as the anterior cerebellum.

**Fig. 3.**
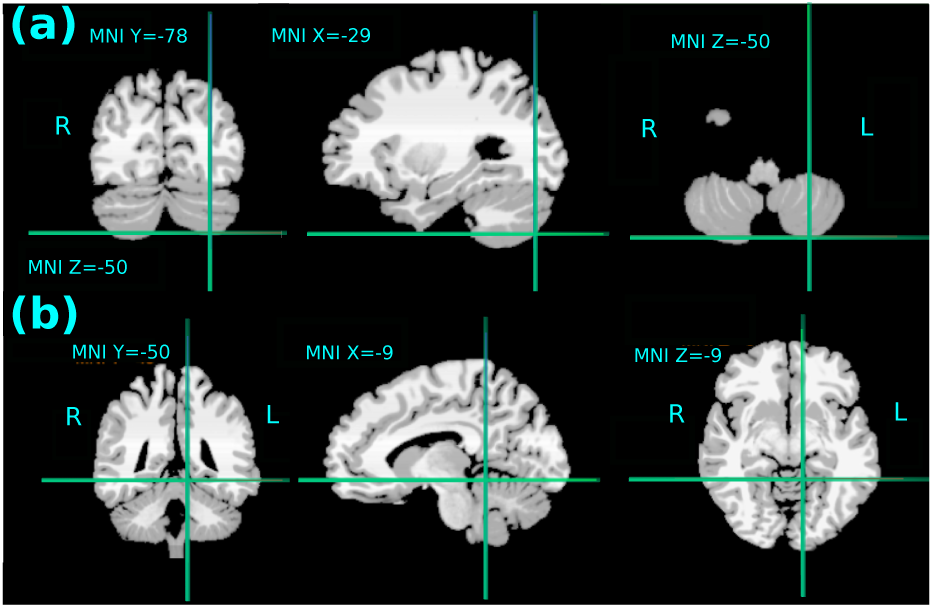
Outside BAS regions involved in the most important correlation, plotted using the Yale BioImage Suite Package web. In (a), the Outside BAS 1 is identified as the anterior cerebellum, while in (b), the Outside BAS 5 is identified as the posterior cerebellum.

Figure 4 summarizes the primary connections identified for each group. Additionally, Table 2.3 provides a structured overview of these connections, where the color intensity reflects their importance ranking, with darker shades indicating a higher significance. In both Figure 4 and Table 2.3, the most relevant connections for ASD, AS, and PDD are highlighted in pink, purple, and green, respectively, ensuring a clear visual distinction between groups.

**Fig. 4.**
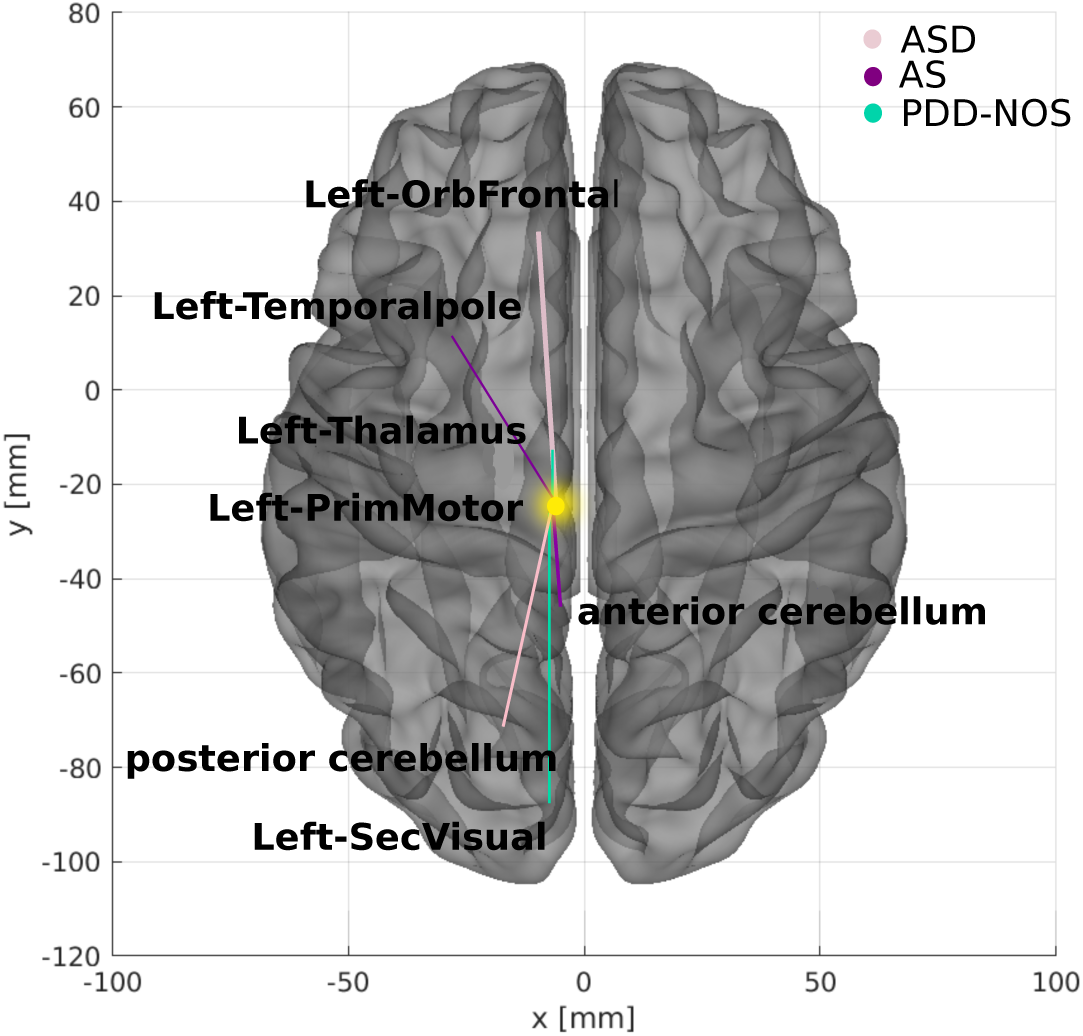
The most important connections identified. A two-dimensional schematic (ventral-axis) highlights the most critical connection for ASD, AS, and PDD-NOS in pink, purple, and green, respectively. The region that most contributed to the primary connection across all groups is highlighted in yellow. The brain plot was generated using the Braph tool [47], with each region mapped according to the Brodmann map from the Yale BioImage Suite Package.

**Table 1.**
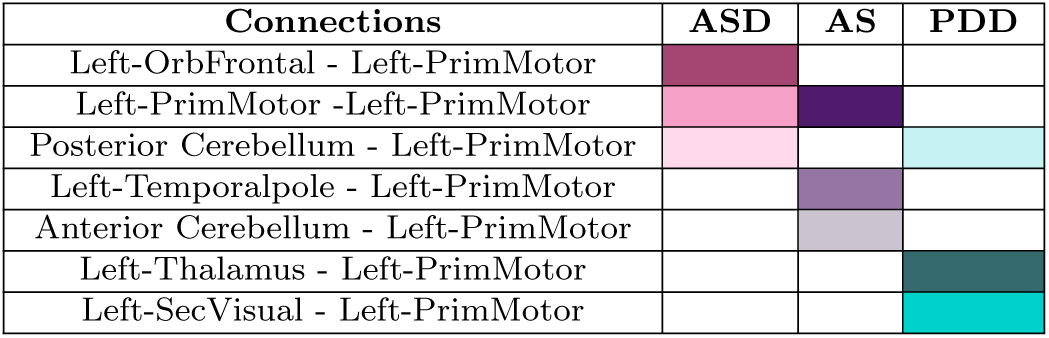
Summary of the most important connections identified for each group using the SHAP value methodology. The connections are categorized by group (ASD, AS, and PDD), with color intensity in each cell representing the importance ranking, with darker shades indicating higher-ranked connections within each group.

**Table 2.**
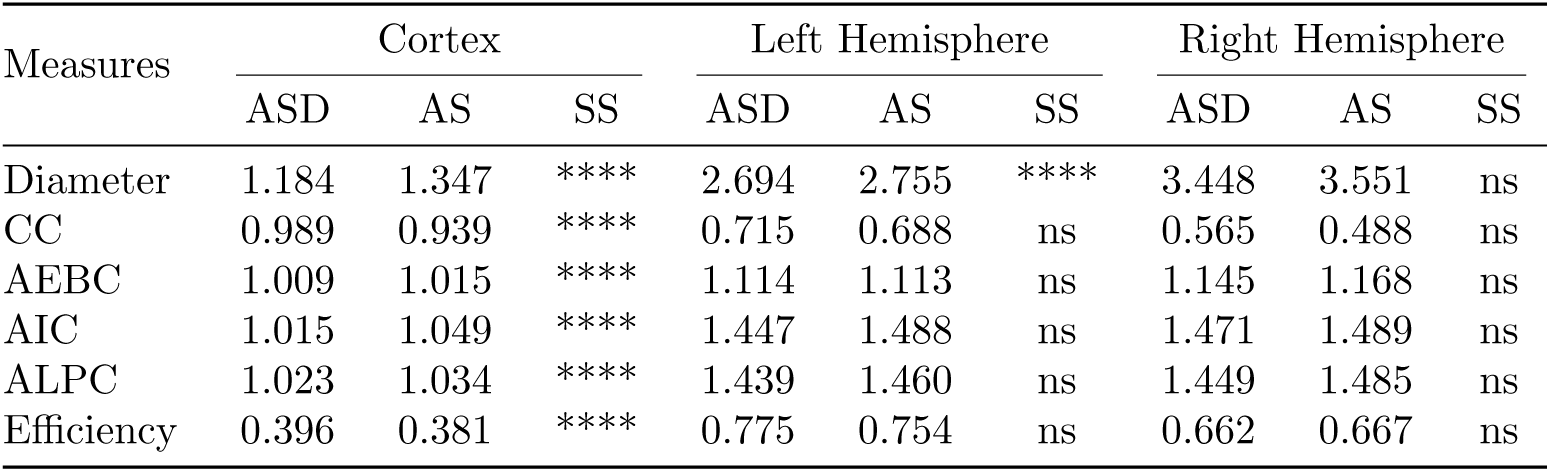
Summary of significant differences in complex network measures between ASD and AS groups across the cortex, left, and right hemispheres. Measures include diameter, CC, AEBC, AIC, ALPC, and efficiency. Statistically significant (SS) differences are marked with stars (****), with the most notable distinctions observed in the cortex. Non-significant differences are indicated as “ns.”

| Measures | Cortex |  |  | Left Hemisphere |  |  | Right Hemisphere |  |  |
| --- | --- | --- | --- | --- | --- | --- | --- | --- | --- |
|  | ASD | AS | SS | ASD | AS | SS | ASD | AS | SS |
| Diameter | 1.184 | 1.347 | **** | 2.694 | 2.755 | **** | 3.448 | 3.551 | ns |
| CC | 0.989 | 0.939 | **** | 0.715 | 0.688 | ns | 0.565 | 0.488 | ns |
| AEBC | 1.009 | 1.015 | **** | 1.114 | 1.113 | ns | 1.145 | 1.168 | ns |
| AIC | 1.015 | 1.049 | **** | 1.447 | 1.488 | ns | 1.471 | 1.489 | ns |
| ALPC | 1.023 | 1.034 | **** | 1.439 | 1.460 | ns | 1.449 | 1.485 | ns |
| Efficiency | 0.396 | 0.381 | **** | 0.775 | 0.754 | ns | 0.662 | 0.667 | ns |

We employed the SHAP values methodology to derive a feature importance vector for each group. To evaluate the similarity between these vectors, we used cosine similarity—a metric that quantifies the directional similarity between two vectors and is widely applied in in the medical literature [48–50]. The cosine similarity scores for each group are visualized in the clustering map presented in Figure 5, which graphically represents the relationships between groups based on their feature importance profiles, facilitating a comprehensive understanding of the clustering patterns.

**Fig. 5.**
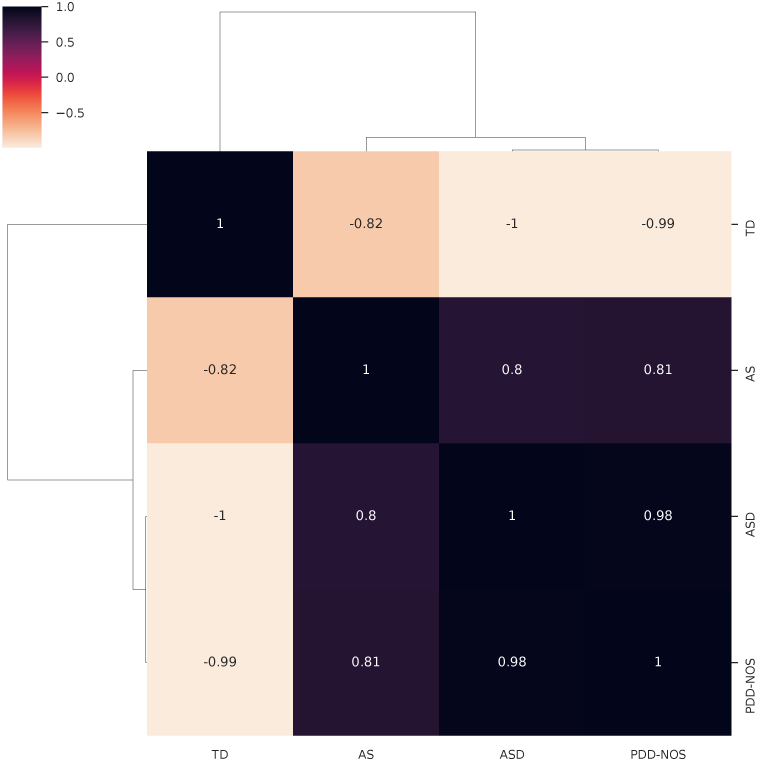
The clustermap visualizes the similarity matrix, with each cell representing the cosine similarity between the SHAP values of two subtypes. The values range from −1 to 1, where 1 indicates identical patterns, 0 implies no similarity, and −1 suggests completely opposite patterns. The off-diagonal elements reveal the degree of similarity between different subtypes.

The resulting connectivity matrix illustrates the pairwise cosine similarity between the SHAP values of different subtypes. The values range from −1 to 1, where 1 indicates identical patterns, 0 implies no similarity, and −1 suggests completely opposite patterns. The off-diagonal elements reveal the degree of similarity between different subtypes. Notably, the TD group exhibits strong negative similarities with ASD, AS, and PDD-NOS, indicating distinct patterns in the identified important connection features. Meanwhile, ASD, Asperger, and PDD-NOS show positive similarities among themselves, suggesting shared connection patterns and potential underlying commonalities. This clustering pattern provides insights into the neural connectivity differences that distinguish individuals with ASD from TD and highlights potential nuances among the ASD subtypes, particularly AS.

Interestingly, the clustering map shows that PDD-NOS shares a 98% similarity with ASD, supporting the hypothesis that PDD-NOS may be a subtype within the broader autism spectrum. Additionally, the AS group shows a comparable level of similarity (approximately 80%) with both PDD-NOS and ASD, suggesting shared features with these groups while still maintaining distinct connection patterns. These clustering patterns provide valuable insights into the neural connectivity profiles identified as important for classification, highlighting the nuanced distinctions among ASD subtypes, particularly in the case of AS, which exhibits both shared and unique connection patterns.

A notable observation from our analysis is that most of the primary regions identified as important by the SHAP values methodology were located in the left hemisphere.

To further explore this hemispheric dominance, we isolated regions corresponding to the left hemisphere (86 regions), right hemisphere (13 regions), and cortex (12 regions). Connectivity matrices were generated separately for each of these subsets and used to train the ML algorithm. However, the classification performance for all subsets remained close to 50%, suggesting that analyzing isolated hemispheres or the cortex independently is insufficient for a robust classification. This limitation likely arises because, in the AS, ASD, and PDD-NOS groups, the most important connections involve cross-hemispheric or more global network patterns. These findings highlight the necessity of incorporating connections across all regions to fully capture the complexity of the neural connectivity patterns that distinguish these groups.

### 2.4 Complex network measures highlight topological alterations in AS

To characterize the network structure of the brain, we computed 26 complex network measurements, as used in our previous publications [51–55]. These measurements include average shortest path length (APL) [56], eccentricity [57], closeness centrality (CC) [58], diameter [59], density [60], betweenness centrality (BC) [61], hub score [62], eigenvector centrality (EC) [63], assortativity coefficient [64, 65], average degree of nearest neighbors [66] (Knn), second moment of the degree distribution (SMD) [67], mean degree [68], entropy of the degree distribution (ED) [69], transitivity [70, 71], complexity, k-core [72], and efficiency [73].

In this study, we employed recently developed metrics (detailed in [52]) that quantify the number of communities within a complex network. Our investigation also incorporated community detection algorithms, as referenced in [74–76]. To integrate these community detection measures into the matrix, which requires a singular scalar value, we applied algorithms designed to identify the largest community. Subsequently, we computed the average path length within each community, resulting in a singular value. The community detection algorithms used in this study included fastgreedy (FC) [77], Infomap (IC) [78], leading eigenvector (LC) [79], label propagation (LPC) [80], edge betweenness (EBC) [81], spinglass (SPC) [82], and multilevel community identification (MC) [83]. o indicate the approach used for calculating the average path length, we extended the respective abbreviations with the letter ”A” (AFC, AIC, ALC, ALPC, AEBC, ASPC, and AMC). Further details of these methodologies can be found in [52].

Within the context of brain networks, two fundamental concepts emerge: integration and segregation. Integration refers to the degree of interconnectedness among nodes in a network, enabling efficient information flow [84, 85]. Highly integrated networks facilitate seamless communication between brain regions [86, 87]. In contrast, segregation reflects the brain’s ability to support specialized processing within tightly interconnected clusters or communities of brain regions [88, 89]. Following our previous research, we employed three measures to analyze these concepts: effective information (*EI*) and determinism and degeneracy coefficients (as elaborated in Alves). Lower *EI* values indicate a more segregated network structure, while higher values of determinism and degeneracy coefficients indicate that the graph structure is characterized by sparse connections rather than a highly interconnected network, further signifying increased segregation.

The performance of the test sample using the complex network approach resulted in a mean AUC of 0.829, precision of 0.938, recall of 0.715, and accuracy of 0.888. However, since the overall model accuracy did not exceed 90%, we opted to prioritize statistical tests over SHAP values for further analysis. Nonetheless, the clear separation of the AS group in the confusion matrix highlights its potential as a distinct subtype within ASD.

The statistical analysis, illustrated in Figure 6, reveals significant differences in network measures among the ASD, AS, and TD groups. Specifically, the AS group consistently exhibited the lowest average values across most metrics, including AFC, ALC, AMC, ASPC, and efficiency, while displaying higher transitivity values compared to TD and ASD. These findings underscore the unique network characteristics of the AS group, as shown in the boxplots, further supporting its distinctiveness within the ASD spectrum.

**Fig. 6.**
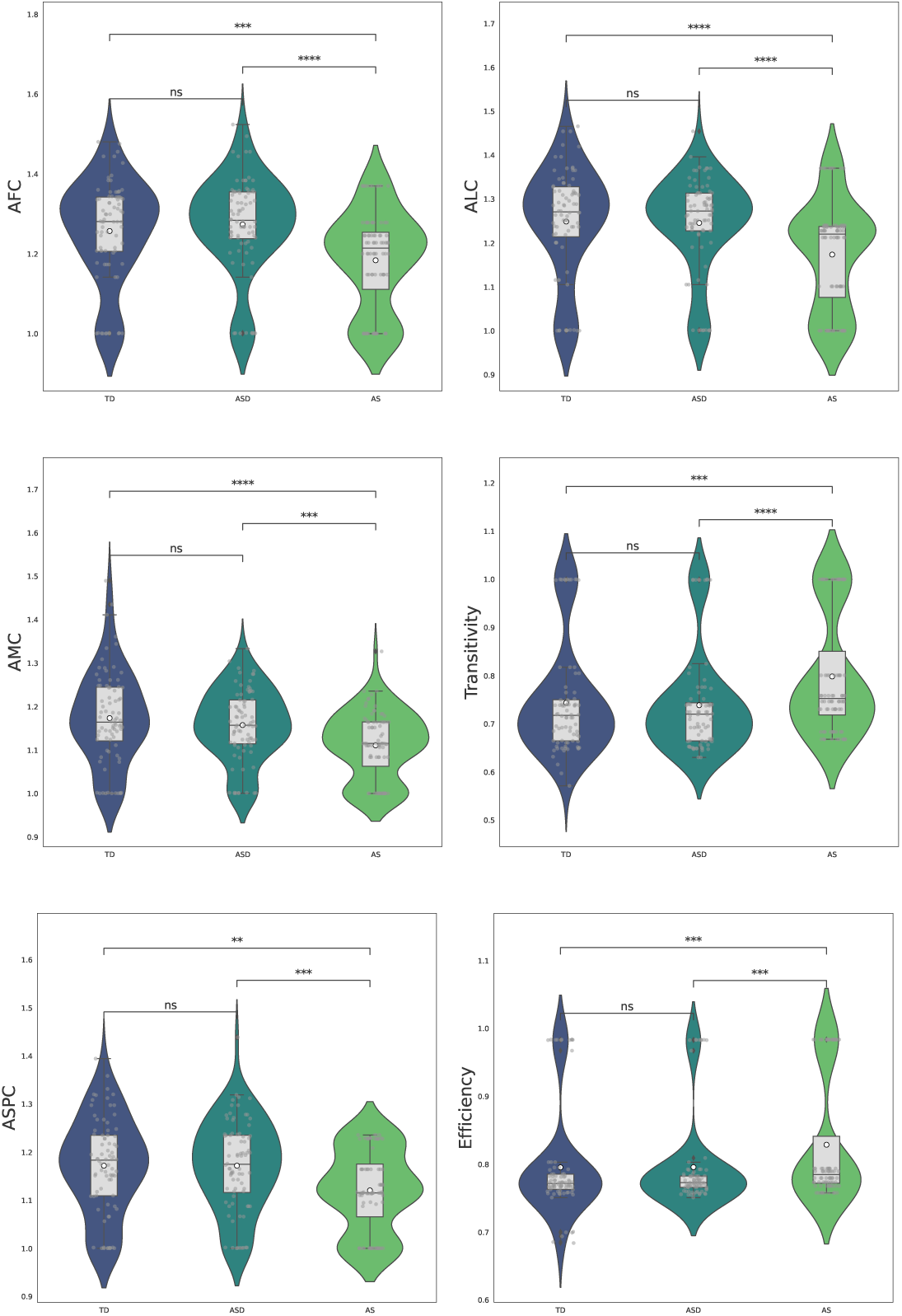
Complex network measures obtained four and three stars in the t-test with Bonferroni correction compared to TD, ASD, and AS classes, represented in dark blue, blue, and green, respectively. Statistical significance is indicated as follows: ns for 5.00 *×* 10*^−^*^2^ *< p ≤* 1.00, * for 1.00 *×* 10*^−^*^2^ *< p ≤* 5.00 *×* 10*^−^*^2^, ** for 1.00 *×* 10*^−^*^3^ *< p ≤* 1.00 *×* 10*^−^*^2^, *** for 1.00 *×* 10*^−^*^4^ *< p ≤* 1.00 *×* 10*^−^*^3^, and **** for *p ≤* 1.00 *×* 10*^−^*^4^.

### 2.5 Unique topological features in AS

In this study, we employed TDA measures, including the number of simplices, fractal dimension, and persistence entropy, to explore the higher-order connectivity of brain networks across ASD, AS, and TD groups. These metrics are further detailed in

Subsection 4.1.3. The statistical analysis, illustrated in Figure 7, reveals significant differences in TDA measures among the three groups. The statistical analysis, visualized in Figure 7, highlights the significant differences in TDA measures among the ASD, AS, and TD groups. The number of simplices was higher in AS compared to TD and ASD, while the fractal dimension followed a similar trend, with AS exhibiting higher values than the other two groups. Persistence entropy, however, exhibited a different pattern, with AS showing the highest values, followed by TD, and ASD exhibiting the lowest. Notably, the most significant differences in TDA metrics were observed when distinguishing AS from TD, with the fractal dimension effectively distinguishing AS from ASD. These findings underscore the distinctiveness of AS as a subtype within the ASD and highlight the value of TDA measures in capturing these differences.

**Fig. 7.**
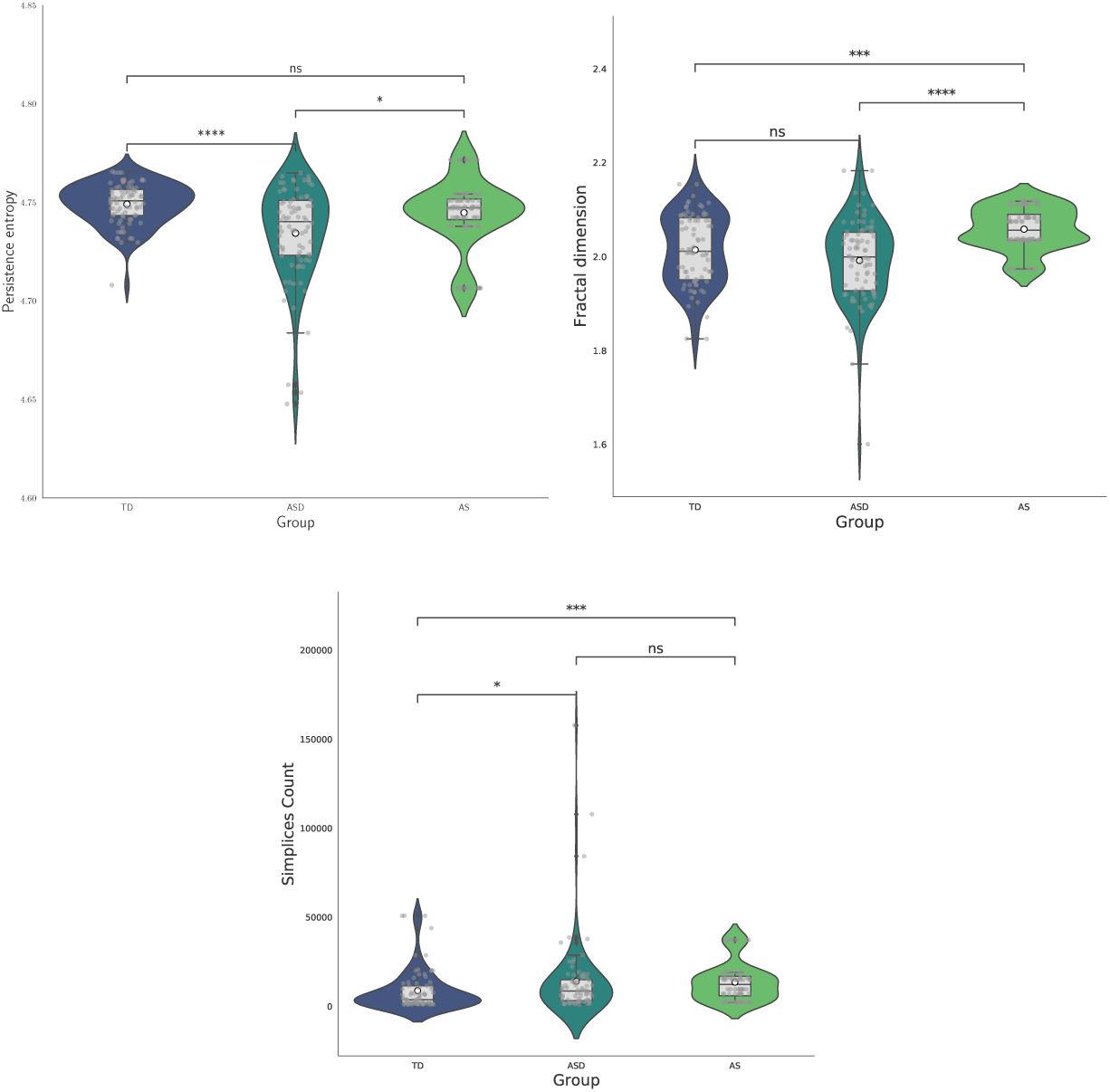
TDA measures obtained four and three stars in the t-test with Bonferroni correction compared to TD, ASD, and AS groups, represented in dark blue, blue, and green, respectively. Statistical significance is defined as follows: ns for 5.00*×*10*^−^*^2^ *< p ≤* 1.00, * for 1.00*×*10*^−^*^2^ *< p ≤* 5.00*×*10*^−^*^2^, ** for 1.00*×*10*^−^*^3^ *< p ≤* 1.00*×*10*^−^*^2^, *** for 1.00*×*10*^−^*^4^ *< p ≤* 1.00*×*10*^−^*^3^, and **** for *p ≤* 1.00*×*10*^−^*^4^.

### 2.6 Hemispheric and regional insights

To gain a deeper understanding of the neural connectivity patterns distinguishing ASD and AS groups, we computed complex network measures for each brain region, focusing on the cortex, left hemisphere, and right hemisphere. While previous analysis showed that isolated connectivity matrices for individual hemispheres or regions were insufficient for robust classification, they underscored the importance of evaluating region-specific topological changes using statistical tests. The results, summarized in Table 2 and visualized in the boxplot in Appendix B, reveal significant differences in several complex network measures across the cortex, left, and right hemispheres. In particular, the cortex’s diameter, CC, AEBC, AIC, ALPC, and efficiency consistently exhibited highly significant differences (****) between ASD and AS groups.

Notably, the diameter measure was higher in AS than ASD across all regions, with the most significant differences observed in the cortex and left hemisphere. CC and efficiency values were generally lower in AS compared to ASD, with significant differences primarily found to the cortex.

The boxplot in Figure 8 provides a detailed comparison of inter- and intra-regional differences between the right and left hemispheres, highlighting the distinct network characteristics of ASD and AS groups. To enhance clarity and focus on the differences between ASD and AS, the TD baseline has been removed from the analysis.

**Fig. 8.**
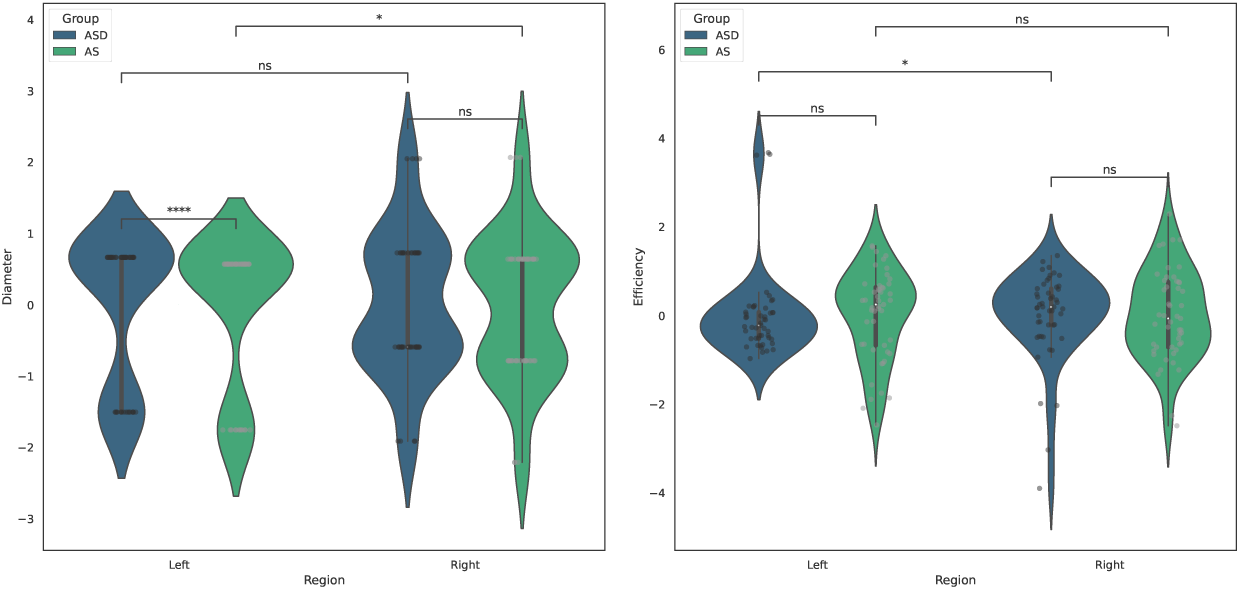
Boxplots illustrating inter- and intra-regional differences in complex network measures (diameter and efficiency) between ASD and AS groups across the left and right hemispheres. Statistical significance is indicated as follows: ns for 5.00 *×* 10*^−^*^2^ *< p ≤* 1.00, * for 1.00 *×* 10*^−^*^2^ *< p ≤* 5.00 *×* 10*^−^*^2^, ** for 1.00 *×* 10*^−^*^3^ *< p ≤* 1.00 *×* 10*^−^*^2^, *** for 1.00 *×* 10*^−^*^4^ *< p ≤* 1.00 *×* 10*^−^*^3^, and **** for *p ≤* 1.00 *×* 10*^−^*^4^.

Among the evaluated metrics, diameter and efficiency showed statistical significance. Inter-regional analysis revealed that the AS group’s left hemisphere had a significantly higher diameter (**) compared to the left hemisphere of the ASD group. Additionally, intra-regional differences were also detected within the AS group, where the left hemisphere’s diameter significantly differed from the right hemisphere. In the ASD group, efficiency showed a notable intra-regional difference, with higher values in the right hemisphere compared to the left. These results highlight both inter-group and hemispheric asymmetries, providing deeper insights into the distinct structural properties of the ASD and AS groups.

## 3 Discussion

### 3.1 Benchmarking ML models for class separation and spatiotemporal correlation patterns

Overall, our approach outperformed existing multiclass ML algorithms for distinguishing ASD, AS, PDD-NOS, and TD, as discussed in Section 1.

The SHAP value analysis provided critical insights into the neural underpinnings of various autism subtypes. Notably, the Left-PrimMotor region consistently emerged as the central hub across all connections, highlighting its crucial role in the neurobiological mechanisms underlying ASD.

In the ASD subtype, the primary connections were identified between the Left-PrimMotor and Left-OrbFrontal regions. Additionally, significant connections involving the posterior cerebellum and Left-PrimMotor. This finding aligns with existing literature emphasizing the role of the frontal lobe, particularly the orbital frontal cortex, in social cognition and executive functions, which are often affected in individuals with ASD [53, 90, 91]. The involvement of the cerebellum is also consistent with studies highlighting its contribution to motor control and cognitive processes, both of which are frequently altered in ASD [92, 93].

For the AS subtype, the principal connections were observed between the Left-PrimMotor and Left-Temporalpole regions. This finding is aligns with existing literature emphasizing the importance of the temporal lobe in processing social information and emotions—functions often impaired in individuals with AS [94, 95]. The involvement of the anterior cerebellum is particularly noteworthy, as research suggests its contribution to higher-order cognitive functions and language processing [93]. In the PDD-NOS subtype, critical connections were identified between the Left-PrimMotor and Left-Thalamus regions, as well as between the Left-SecVisual and Left-PrimMotor. The thalamus plays a well-documented role in sensory processing and integration, and its involvement in PDD-NOS further underscores the heterogeneity within the autism spectrum [96, 97]. Additionally, the involvement of the secondary visual cortex may indicate variations in visual processing, an aspect frequently reported in individuals with PDD-NOS [98].

Notably, the cluster map derived from the SHAP value methodology suggests that PDD-NOS is closely related to ASD, with a cosine similarity of 0.98. In contrast, the similarity between ASD and AS is 0.81. Since this value is lower than 0.90, it indicates that AS exhibits more distinct characteristics compared to ASD. This finding suggests that AS patients should be analyzed separately from the ASD group rather than being grouped within the broader ASD category.

### 3.2 Complex network measures highlight topological alterations in AS

The statistical analysis reveals significant differences in network measures among ASD, AS, and TD groups, with AS group uniquely exhibiting higher values for both global average efficiency and global transitivity. In contrast, TD and ASD groups showed no distinguishable differences in these metrics, underscoring the distinctiveness of AS group within the ASD spectrum. Furthermore, the AS group consistently displayed the lowest average values across most metrics, including AFC, ALC, AMC, and ASPC, while maintaining higher values for efficiency and transitivity.

Global average efficiency, which quantifies the ease and speed of information transfer across the entire network [99, 100], was notably higher in the AS group. This suggests that individuals with AS may exhibit enhanced global integration, facilitating more effective communication between distant brain regions. Similarly, global transitivity, defined as the ratio of triangles to connected triples in the network [101, 102], was also higher in the AS group, indicating strong local clustering and a well-defined modular organization. These findings highlight a unique balance between global and local network properties in the AS group, differentiating it from both ASD and TD groups.

In contrast, the consistently lower values of metrics such as AFC, ALC, AMC, and ASPC in AS group suggest a more compact and efficient internal structure within communities. This pattern may indicate that the AS group prioritizes tightly connected local modules while simultaneously maintaining effective global integration. Such a network configuration could facilitate enhanced cognitive abilities and localized processing capabilities, characteristics often associated with AS, which is frequently described as a high-functioning form of ASD [103–105].

The higher efficiency and transitivity values in the AS group align with characteristics traditionally associated with this syndrome, including stronger cognitive abilities and better social functioning compared to others on the ASD spectrum [106–108]. The combination of enhanced global integration and robust local clustering may contribute to the cognitive flexibility and social abilities observed in individuals with AS, despite challenges in other areas. Our findings are consistent with those of[109], which also reported increased global efficiency in AS compared to ASD and TD. Moreover, the absence of significant differences in efficiency and transitivity between TD and ASD groups further underscores the distinctiveness of AS group.

### 3.3 Unique topological features in AS

The TDA results further complement the complex network findings, highlighting the distinctiveness of AS group within ASD. Statistical analysis of TDA metrics revealed significant differences among ASD, AS, and TD groups, with AS group displaying unique topological characteristics. Specifically, AS group exhibited higher values for the number of simplices and fractal dimension, indicating greater network complexity and hierarchical organization. The high value of fractal dimension is consistent with the increased global efficiency and transitivity observed in the complex network analysis, suggesting a more intricate and hierarchically organized network with robust local clustering. The high value of fractal dimension in AS may also correlate with the cognitive and behavioral traits associated with high-functioning ASD, implying that such intricate network structure could underlie their superior verbal skills and cognitive flexibility compared to other ASD subtypes.

Persistence entropy provided an additional dimension of analysis by quantifying the variability and prominence of topological features, such as connected components and loops, across multiple thresholds. Interestingly, persistence entropy exhibited the highest values in AS group, followed by TD, with ASD showing the lowest values. These findings suggest that AS networks possess a more complex and variable topology, potentially reflecting a combination of flexibility and intricate neural connectivity that may underlie the group’s distinct cognitive and behavioral traits. The higher persistence entropy in AS may indicate an improved ability to integrate and process different information streams. In contrast, the lower persistence entropy in ASD suggests a more constrained and less adaptable network organization, potentially contributing to the group’s characteristic difficulties in information processing and social interaction. The intermediate persistence entropy observed in TD reflects a balanced and flexible network structure, supporting a diverse range of neural functions.

The distinctive combination of higher fractal dimension and lower persistence entropy in AS group underscores a balance between network complexity and specialization. These findings provide valuable insights into how hierarchical organization and topological variability differentiate AS from other groups within the ASD spectrum. Moreover, the ability of persistence entropy to capture subtle differences between ASD and AS subtypes highlights its potential as a powerful metric for understanding neurodevelopmental disorders.

To the best of our knowledge, this is the first study to apply TDA metrics, including persistence entropy, to investigate brain network topology in AS and ASD. These findings highlight the potential of TDA to uncover novel insights into the neurobiological underpinnings of ASD subtypes, particularly AS, and pave the way for future research on targeted interventions and diagnostic tools.

### 3.4 Hemispheric and regional insights

The results reveal significant differences in network topology between ASD and AS groups, particularly in global efficiency, CC, and community detection metrics, as summarized in Table 2. Moreover, these differences highlight distinct patterns of integration and modularity at both regional and global levels ((discussed in Subsections 2.4 and 2.6, respectively). The AS group exhibits pronounced hemispheric asymmetries and localized inefficiencies, especially in the right hemisphere, while also displaying higher global network efficiency and transitivity compared to the ASD group. At the regional level, AS networks show pronounced hemispheric asymmetries, with the right hemisphere characterized by a larger diameter, lower CC, and reduced efficiency compared to the left hemisphere. his suggests fragmented integration in localized networks. These findings align with previous research indicating reduced right-hemisphere function in AS, which may contribute to difficulties in social communication and spatial processing [110, 111].

In contrast, the ASD group exhibits less pronounced hemispheric differences, with both hemispheres maintaining relatively high efficiency and similar CC values. This aligns with reports of reduced lateralization in ASD [112–114]. However, at the global level, AS networks exhibit higher global average efficiency and global transitivity compared to ASD, reflecting greater global integration and modularity. These findings are consistent with previous research suggesting increased global efficiency in AS, which may reflect improved long-range connectivity and more effective information transfer across the brain [108, 115]. This unique combination of regional inefficiencies and global robustness could explain some of the clinical characteristics traditionally associated with AS as a high-functioning form of autism. The higher global efficiency and transitivity in AS, also supported by the literature [109] could contribute to advanced cognitive functions, such as problem-solving, language processing, and analytical reasoning. Meanwhile, localized disruptions in the right hemisphere may be related to interpersonal difficulties [116, 117]. These network properties could underpin the superior verbal skills and intellectual capabilities often observed in individuals with AS, despite challenges in social interactions and emotional processing [118, 119].

The pronounced hemispheric asymmetries in AS, particularly the reduced integration in the right hemisphere, may reflect functional differences associated with cognitive and behavioral traits. The right hemisphere, often associated with spatial and emotional processing, shows reduced integration in AS, which could contribute to the interpersonal difficulties commonly reported in this group [120, 121]. Conversely, the relatively preserved integration and efficiency in the left hemisphere may support the advanced language and reasoning abilities characteristic of high-functioning ASD [122].

### 3.5 Summary of distinct neural profiles of ASD subtypes: insights, implications, and limitations

This study provides new insights into the neurobiological differences between ASD and AS, highlighting distinct patterns of intra-brain correlations and network topology across different scales. Notably, the AS group exhibits pronounced hemispheric asymmetries, with reduced integration and efficiency in the right hemisphere compared to the lef, while also demonstrating higher global efficiency and transitivity than ASD. These findings suggest a unique balance between global robustness and regional fragmentation in AS, which may contribute to its classification as high-functioning autism. The application of TDA metrics, including fractal dimension and Betti numbers, further enhances this understanding, as AS networks display a higher fractal dimension and fewer Betti numbers, indicating greater complexity and stronger integration. This study is the first to systematically compare the right, left, and cortical hemispheres of ASD and AS while integrating TDA, providing a novel framework for investigating brain connectivity. These findings support the broader perspective of AS as an distinct subtype within ASD spectrum.

While this study provides valuable insights into the neurobiological differences between ASD and AS, several limitations should be acknowledged. First, small sample size, particularly for certain subtypes such as PDD-NOS, restricts the statistical power and generalization of the findings. Additionally, although hemispheric and cortical analysis revealed important distinctions, the study does not account for potential confounding factors such as age, sex, or comorbidity, which may also influence brain network properties. Future research with larger longitudinal datasets and more detailed assessments of clinical phenotypes is necessary to validate and further develop these findings.

## 4 Methods

### 4.1 Data and data preprocessing

The dataset used for this analysis, as in our previous work [53], originates from the ABIDE consortium, which comprises 1,112 datasets, including 539 individuals with ASD and 573 TD controls. The preprocessed data was obtained from the Preprocessed Connectomes Project (PCP) dataset, where a rigorous preprocessing pipeline was applied. This pipeline included slice-time correction, motion correction, intensity normalization, and removal of various artifacts such as breathing, heartbeat, and head motion. For each subject, a 300-second BOLD time series was used.

All data were strictly anonymous in compliance with HIPAA requirements, and analyses were conducted following the pre-approved protocols of the University of Utah Institutional Review Board. Informed consent was obtained from all participants according to the established procedures of human subjects research committees at each participating institution. Notably, the preprocessed data underwent a 0.5 Hz band-pass filter, consistent with recent fMRI literature findings indicating the relevance of fluctuations within this frequency range. Detailed information on data acquisition, informed consent procedures, and site-specific protocols can be accessed at http://fcon1000.projects.nitrc.org/indi/abide/. Moreover, the dataset is available for analysis using the Nilearn python package [19], a widely used tool for neuroimaging data processing and exploration, cited in numerous studies [123–127].

In this study, we extend our previous work [53] by providing a more detailed perspective on ASD through the inclusion of four distinct diagnostic groups: AS (N=80), PDD-NOS (N=48), ASD (N=282), and TD (N=518). Further, this work focuses on specific brain regions rather than analyzing the entire BOLD time series from each voxel in the brain image. Among various predefined atlases, we selected the Bootstrap Analysis of Stable Clusters (BASC) due to its superior performance in distinguishing individuals with ASD using a deep learning model, as reported in [53, 128]. Originally introduced by [129], the BASC atlas is derived from group-level brain parcellation using the BASC method, a k-means clustering algorithm that identifies brain networks with coherent activity during resting-state functional Magnetic Resonance Imaging (fMRI) [130, 131]. The BASC atlas used in this study consists of 122 regions of interest (ROIs) (see Figure 9-A). In our previous research [53], we used the Yale BioImage Suite Package web application^1^ to manually assign coordinates to each ROI and determine their anatomical labels (see Figure 9-A).

**Fig. 9.**
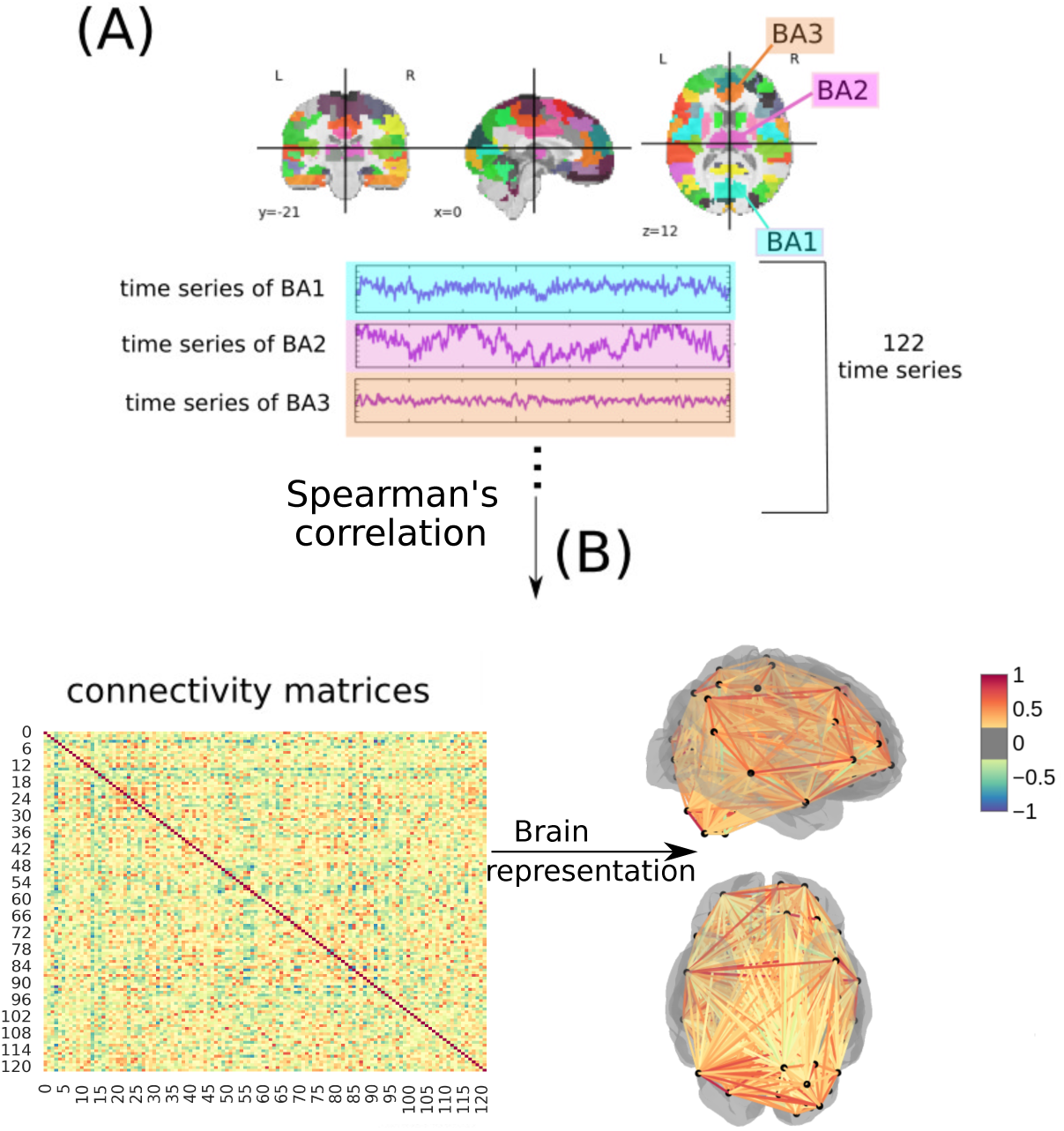
Approach for generating connectivity matrices as described in. [53]. In (A), a time series is extracted from 122 ROIs in the fMRI data using the BASC BOLD atlas (highlighted in blue, purple, and orange). These time series are then correlated (B) to generate the connectivity matrices, where each row and column corresponds to one of the Brodmann areas for a patient with AS, ASD, PDD-NOS, or TD (the figure illustrates an example for a subject with AS). The highlighted matrices represent the brain, as shown in a three-dimensional scheme (from bottom and right perspectives) .

After extracting time series data for each patient across 122 regions, we performed correlation analysis using Spearman’s correlation coefficient [132] (see Figure 9-B).

This method was selected based on its superior performance in our previous study [53], where it effectively differentiated ASD patients from TD controls. Spearman’s correlation measures the monotonic relationship between variables, making it a robust metric for assessing associations, particularly in the presence of nonlinear dependencies within brain data [133–135].

#### 4.1.1 ML models pipeline

First, we applied standardization, a widely used ML technique to normalize features setting the mean to zero and the standard deviation to one [136]. This transformation is crucial for simplifying data analysis, ensuring consistent scaling, equal weighting of features, and increasing resilience to outliers and highly variable features within the model. Subsequently, we split the data into training and test sets, with 30% of the data comprising the test set, a common practice in the literature [137–140].

To ensure robustness and unbiased evaluation of machine learning models in the training set, we consistently applied 10-fold stratified cross-validation with shuffling [141, 142]. This technique separates the dataset into ten equally stratified folds, ensuring that each fold (denoted as *k*) maintains a balanced distribution of samples across all classes. This approach guarantees equal representation of each class in every fold, enhancing the reliability of model evaluations. The algorithm trains on nine folds and validates on the remaining one, repeating this process ten times, with each fold serving as the validation set once. We used the common value of *k* = 10, as recommended in the literature [143–146]. Furthermore, to assess the stability of the model’s performance, we also explored other values of *k*. The shuffling strategy ensures tthat the data is randomized before splitting into folds, mitigating potential ordering effects and biases, thereby strengthening the robustness of the model training process [147].

We employed hyperparameter optimization techniques to achieve optimal performance of the ML algorithm. The grid search method, widely adopted in the literature [41, 148–151], was used for fine-tuning hyperparameters in all ML algorithms, except for CNN. This method involves systematically searching through a predefined set of hyperparameter values to find the combination that yields the best model performance [152]. For deep learning models, specifically CNNs, we opted for random search optimization due to its superior computational efficiency in hyperparameter tuning compared to grid search. This choice was motivated by its superior computational efficiency in hyperparameter tuning compared to grid search. This is especially advantageous considering the substantial computational resources required for deep learning tasks. Detailed information on the hyperparameter optimization values for each classifier model can be found in [51–54].

In evaluating the stability of the model across different values of *k* in stratified cross-validation combined with SVM, as shown in Figure 10, we observed variations in both training and test performance metrics. The training AUC consistently increased with higher values of *k*, reaching a peak at *k* = 10 before slightly decreasing. Similarly, the training accuracy steadily improved with *k*, stabilizing around *k* = 7 and maintaining a high level thereafter. The overall stability of the model is supported by the relatively small fluctuations in test metrics and the convergence of training metrics, particularly after *k* = 7. Despite some variability, the model consistently achieves high performance, indicating that the chosen configuration provides robust results for the dataset and classification task at hand.

**Fig. 10.**
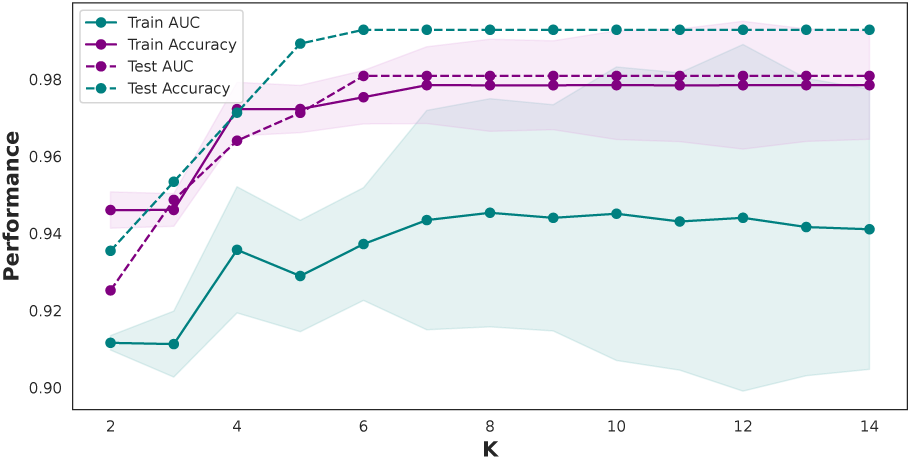
SVM model performance varying the number of *K* in stratified cross-validation. The figure illustrates the performance metrics of the SVM model, specifically AUC and accuracy, represented in blue and purple on the y-axis. These metrics were obtained by varying the number of *K* in the stratified k-fold cross-validation, shown on the x-axis. Dashed lines represent the results from the test set, while solid lines represent the outcomes from the training set. The shaded region indicates the standard deviation within the training set.

To identify the optimal number of features required to achieve peak performance, we conducted a Recursive Feature Elimination (RFE) analysis, as shown in Figure 11. This method, widely used in predictive modeling, particularly in medical data applications [153–156], systematically removes less influential features in a stepwise manner to evaluate their impact on model performance. Through this iterative process, we identified the most relevant features. As shown in Figure 11, the highest accuracy is achieved with a set of 176 features. This finding indicates that using the entire feature set is unnecessary for optimal performance.

**Fig. 11.**
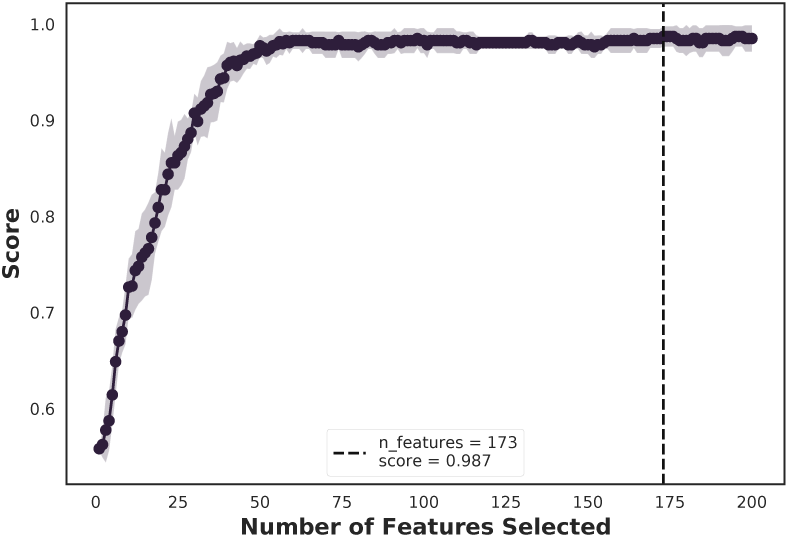
RFE. Optimal performance is achieved with 176 features, indicating that the full set of features is unnecessary for achieving peak efficacy.

Subsequently, we applied the SHAP value approach to provide a biological interpretation, highlighting the predictive significance of individual attributes.

#### 4.1.2 Statistical test

Finally, a statistical analysis was conducted to compare the four distinct groups: ASD, AS, PDD-NOS, and TD. Due to the varying group sizes, random selection was employed, with the minimum number of individuals in any group set at 48, consistent with the PDD-NOS group.

The Wilcoxon test, also known as the Mann-Whitney U test for two groups or the Kruskal-Wallis test for more than two groups [157–159], is a non-parametric test used in this study to compare two groups. Specifically, the Wilcoxon test was employed to identify significant differences in measures or variables among the ASD, AS, PDD-NOS, and TD groups.

Furthermore, we applied the Bonferroni correction to address the potential issue of inflated Type I error rates (false positives), which occur when a null hypothesis that is actually true is incorrectly rejected in a statistical test [160]. This correction was used in our study to control for the increased risk of Type I errors arising from conducting multiple simultaneous statistical tests [161].

The following symbols represent the statistical significance:

- ns: 5.00*e −* 02 *< p <*= 1.00*e* + 00
- *: 1.00*e −* 02 *< p <*= 5.00*e −* 02
- **: 1.00*e −* 03 *< p <*= 1.00*e −* 02
- ***: 1.00*e −* 04 *< p <*= 1.00*e −* 03
- ****: *p <*= 1.00*e −* 04

#### 4.1.3 TDA metrics

In this study, we employed Vietoris–Rips complex, also known as the Rips complex, a fundamental tool in TDA, to investigate the higher-order connectivity of brain networks across ASD, AS, and TD groups [162, 163]. Rips complexes represent the connectivity of data points as simplices (points, edges, triangles, and higher-dimensional structures) based on a specified distance threshold, capturing the underlying topological features of a dataset [164, 165]. Using the Gudhi library [166], we computed the number of simplices up to a fixed maximum dimension (*d*_max_ = 2), capturing points, edges, and triangular structures [167]. For each group, correlation matrices were used to construct Rips complexes, applying a distance threshold of 0.5 to retain only the strongest connections, thereby emphasizing the core connectivity structure of the networks [168]. This metric reflects the density of higher-order connectivity in the networks and provides insights into the underlying neural architecture [169].

We employed the fractal dimension to quantify the complexity and hierarchical organization of brain networks by analyzing the growth of simplices in Rips complexes across multiple thresholds (0.1 to 1.0) and dimensions (*d*_max_ = 1 to 4) [170]. This method captures the scaling behavior of brain connectivity, revealing how topological features emerge at varying connectivity thresholds [171]. The fractal dimension was calculated as the slope of the log-log relationship between the number of simplices and the thresholds, providing a quantitative measure of the network’s self-similarity and multi-scale organization [172]. Lower dimensions (*d*_max_ = 1) capture basic connectivity patterns, while higher dimensions (*d*_max_ = 2, 3, 4) represent more intricate features such as loops and voids, indicative of complex network interactions.

In this study, persistence entropy, derived from persistent homology, quantifies the variability of topological features, such as connected components and loops, as they persist across multiple thresholds in Rips complexes. [20]. Using the Gudhi library, we calculated persistence entropy from Rips complexes and persistence diagrams, capturing the variability of these topological features across thresholds ranging from 0.1 to 1.0. This measure effectively captures the distribution and lifespan of topological features [20, 173–175].

## Data Availability

All data produced in the present study are available upon reasonable request to the authors.

## Footnotes

1 Accessible at https://bioimagesuiteweb.github.io/webapp/mni2tal.html

## Acknowledgements

We thank Marius Oechsner for his insightful suggestions on brain lateralization analysis, particularly the distinctions between left and right hemispheres, and his invaluable support throughout this work.

## A SHAP values plot

**Fig. 12.**
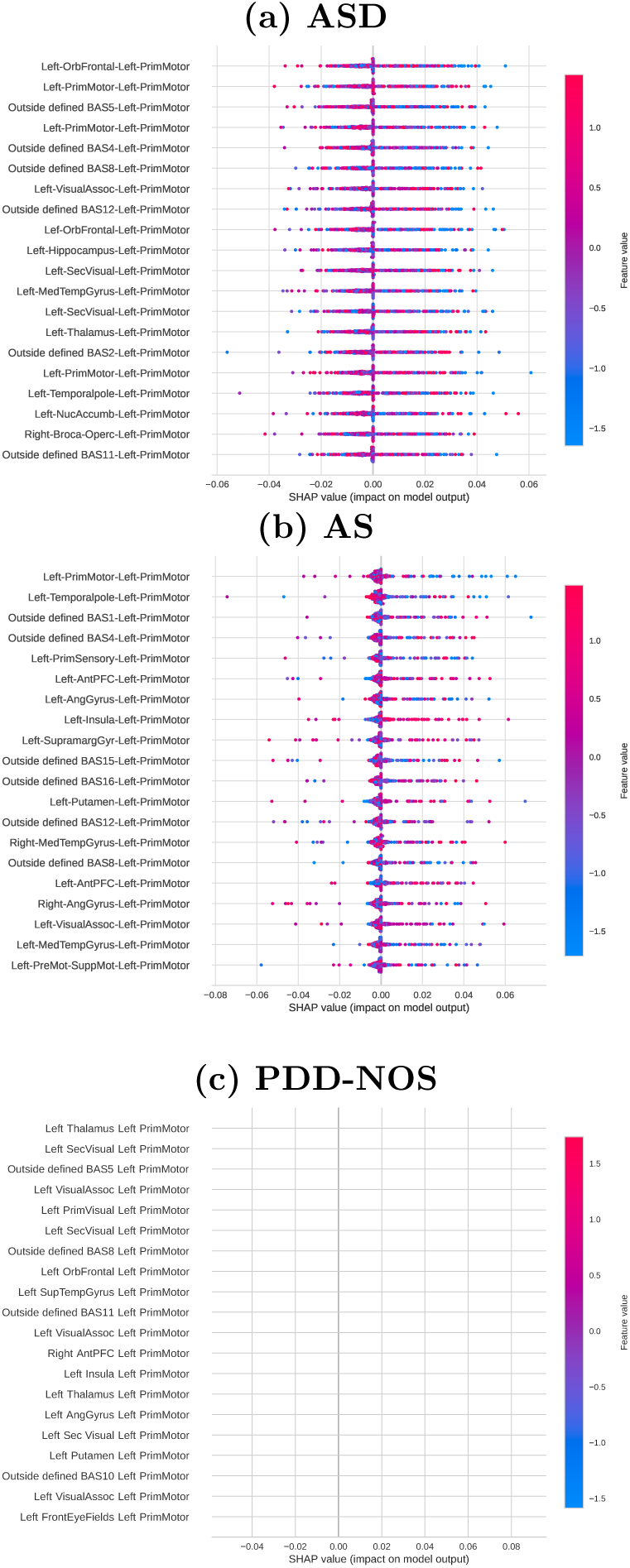
Feature importance ranking using the SHAP values methodology for the SVM classifier, with brain regions in descending order: (a) Feature importance ranking for the ASD class. (b) Feature importance ranking for the AS class. (c) Feature importance ranking for the PDD-NOS class.

## B Complex Network Measures Across Regions

The boxplot figures in 13 illustrate the statistical differences in complex network measures between ASD and AS groups across the cortex, left, and right hemispheres. To facilitate comparisons and emphasize distinctions between ASD and AS, the TD baseline has been excluded from these figures. This adjustment enables a more focused analysis of the specific differences between ASD and AS groups. The figures visually support the summarized results in Table 2, highlighting regional differences in measures such as diameter, CC, AEBC, AIC, ALPC, and efficiency. Specifically, the figures show higher diameter values for AS compared to ASD across all regions, as well as lower CC and efficiency values for AS compared to ASD in the cortex. Non-significant differences (ns) in measures such as CC and efficiency in the left and right hemispheres further emphasize the localized nature of these distinctions in the cortex.

**Fig. 13.**
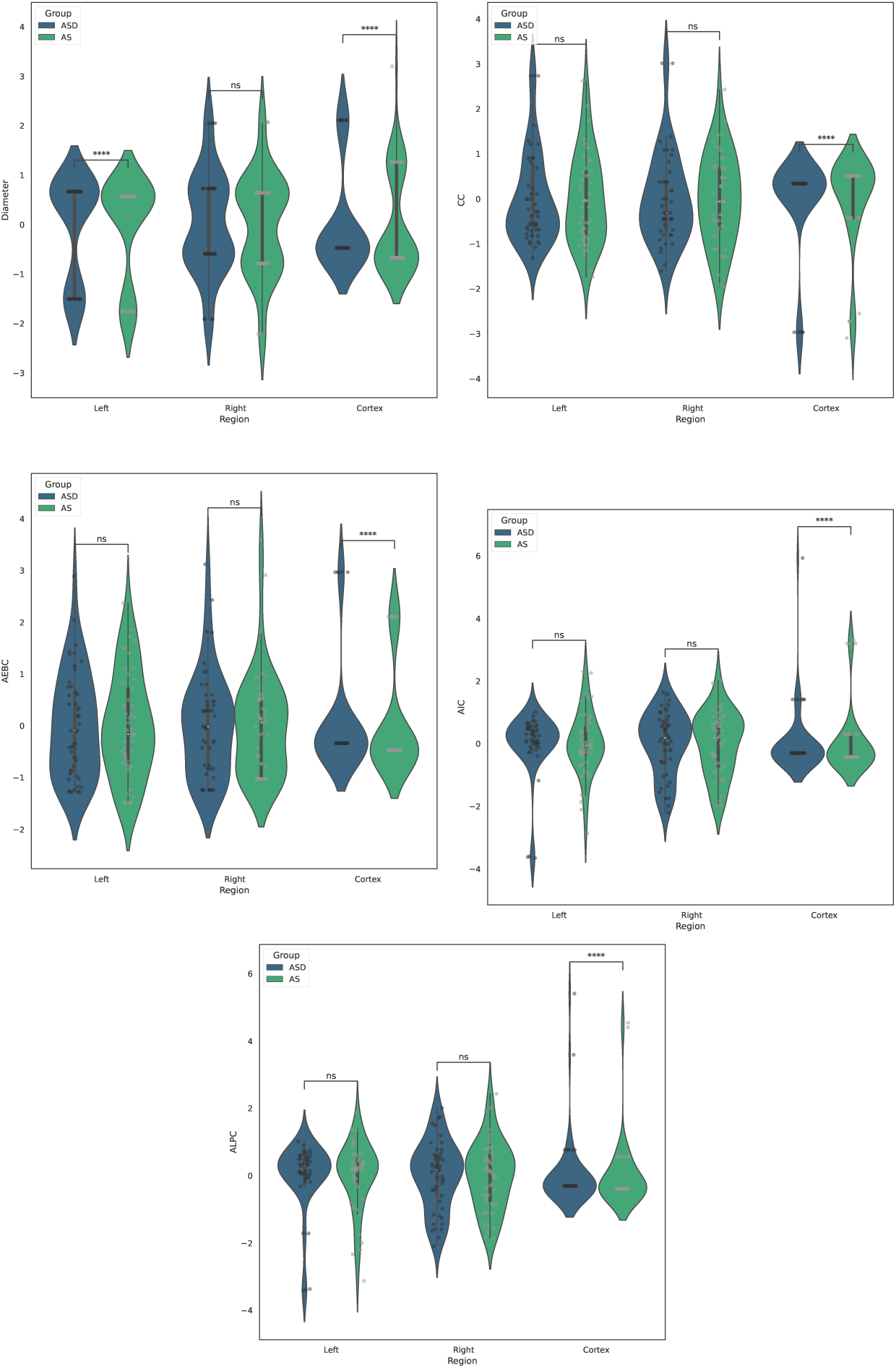
Complex network measures that achieved four and three stars in the t-test with Bonferroni correction are compared across the TD, ASD, and AS classes, represented by dark blue, blue, and green, respectively. Statistical significance is indicated as follows: ns for 5.00 *×* 10*^−^*^2^ *< p ≤* 1.00, * for 1.00 *×* 10*^−^*^2^ *< p ≤* 5.00 *×* 10*^−^*^2^, ** for 1.00 *×* 10*^−^*^3^ *< p ≤* 1.00 *×* 10*^−^*^2^, *** for 1.00 *×* 10*^−^*^4^ *< p ≤* 1.00 *×* 10*^−^*^3^, and **** for *p ≤* 1.00 *×* 10*^−^*^4^.

